# Structural, functional, and quality aspects of social connections and their associations with anthropometric, cardiovascular, and metabolic markers of health in Understanding Society

**DOI:** 10.64898/2026.09.14.26363022

**Authors:** Saoirse Finn, Ellie Roberts, Katie Taylor, Daisy Fancourt

**Affiliations:** Department of Behavioural Science and Health, University College London; Department of Psychology and Human Development, University College London; Department of Epidemiology and Public Health, University College London

**Keywords:** social connections, social support, living alone, relationship strain, cardiovascular, metabolic, adiposity, biomarkers

## Abstract

Social connections influence cardiovascular and metabolic outcomes, but more research is needed to examine whether different aspects of social connections relate differently to physical, physiological, and biological markers of cardiometabolic health. This study examined associations between structural, functional, and quality aspects of social connections and key anthropometric and cardiometabolic markers. Using data from Understanding Society, linear regression analyses adjusted for identified confounders explored associations between living with others (structural), social support (functional), and relationship strain (quality) with anthropometric (BMI, waist circumference; n=16,284), cardiovascular (pulse rate, systolic blood pressure (BP), diastolic BP; n=13,902), and metabolic markers (triglycerides, total cholesterol, glycated haemoglobin (HbA1c); n=10,070). Living with others was associated with higher BMI (B=0.638, 95%CI=0.365 to 0.911) and waist circumference (B = 1.597, 95%CI = 0.928 to 2.265), while greater social support and lower relationship strain showed the reverse association. Living with others, greater social support, and lower relationship strain were all associated with lower pulse rate, although just social support and low relationship strain were associated with lower diastolic BP and none of the three exposures with lower systolic BP. Greater social support and lower relationship strain (but not living with others) were associated with lower total cholesterol (B = - 0.019, 95%CI = -0.033 to -0.005; B = -0.022, 95%CI = -0.039 to -0.006), triglycerides (B = -0.012, 95%CI = -0.018 to -0.005; B = -0.014, 95% CI = -0.022 to -0.006), and HbA1c levels (B = -0.003, 95%CI = -0.005 to -0.001; B = -0.006, 95% CI = -0.008 to -0.003). There was some suggestion of moderation by sex, although this was modest. These findings enhance understanding of the nuanced relationship between structural, functional, and quality aspects of social connections and health.

## Introduction

Social connections play an important role in individuals’ health and wellbeing (Hawkley & Cacioppo, 2010; Holt-Lunstad, 2024). One of the well-established frameworks of social connections categorises relationships into structural, functional, and quality indicators (Holt-Lunstad et al., 2017). Structural aspects refer to the existence and roles of social connections, such as marital status and whether individuals are socially isolated. Functional indicators reflect the actual or perceived availability of aid or resources provided by relationships, including social support. Quality indicators capture the positive and negative aspects of social connections, including whether there is strain within relationships (Holt-Lunstad, 2018). Deficits in social connections, such as social isolation, living alone, loneliness, lack of social support and high relationship strain, have been well-evidenced to have numerous detrimental effects on physical health, including increased risk of cardiometabolic disorders and mortality. For example, meta-analyses and longitudinal studies have shown that social deficits are linked with higher risk of stroke, coronary heart disease, and Type 2 diabetes (Albasheer et al., 2024; Asif et al., 2025; T. W. Smith, 2022; Valtorta et al., 2016), as well as cardiovascular-related and all-cause mortality (Bookwala & Gaugler, 2020; Bulanda et al., 2016; Paul et al., 2021; F. Wang et al., 2023). However, protective effects of social connections have also been evidenced. For example, being more socially engaged, such as through volunteering, has been linked to lower risk of incident cardiovascular disease (CVD) and heart attack (Bell & Ferraro, 2025; Han et al., 2017), as well as lower levels of CVD risk factors, such as diastolic and systolic blood pressure and cholesterol (Kim et al., 2023; Schreier et al., 2013).

Some of the proposed mechanisms that may link social connections to cardiometabolic outcomes involve physiological and biological processes across immune, neuroendocrine, cardiovascular, and metabolic systems (Paul et al., 2021). For example, social deficits, such as greater loneliness and family relationship strain, have been related to higher levels of inflammatory activity (Gao et al., 2024; K. J. Smith et al., 2020; Yang et al., 2014). Low social support has been associated with higher systolic blood pressure (Yang et al., 2015) and higher body mass index (BMI) (Tymoszuk et al., 2019). Being more socially isolated has been associated with both lower systolic and diastolic blood pressure (Shankar et al., 2011). Furthermore, greater frequency of social interactions has been associated with lower levels of blood glucose and low-density lipoprotein (LDL) cholesterol (‘bad’ cholesterol) (Floyd et al., 2017), and greater social support has been related to lower HbA1c levels (Smalls et al., 2022), as well as higher levels of high-density lipoprotein (HDL) cholesterol (‘good’ cholesterol) (Fischer Aggarwal et al., 2008).

However, there remain a number of limitations in this literature. First, research assessing anthropometric and cardiometabolic markers has often focused on a singular marker or a limited subset of markers, rather than examining a broader profile. Second, studies investigating biomarkers indicative of metabolic function, such as HbA1c, triglycerides, and cholesterol, have often been conducted in specific clinical populations or smaller experimental samples, rather than large population-based cohorts. For example, many social connections and cardiovascular marker studies have been undertaken alongside laboratory stress tests or in relation to occupational experiences. Third, studies have typically focused on only one or two aspects of social connections at a time, limiting consideration of whether structural, functional, and quality aspects show distinct associations with these markers.

Therefore, the current study utilised a well-established framework of social connections to investigate the associations between structural, functional, and quality aspects, with a panel of anthropometric, cardiovascular, and metabolic markers of health in a UK cohort study. Addressing this gap is important because identifying associations between different aspects of social connections and broader biomarker profiles could improve mechanistic understanding of how social connections influence leading causes of disability and mortality, including cardiovascular disease and diabetes.

## Methods

### Dataset

Data were drawn from Understanding Society; a longitudinal panel survey of approximately 40,000 UK households, with data currently available across fifteen annual waves collected since 2009 (Institute for Social and Economic Research. University of Essex, 2024). Between 2010 and 2012, adult participants (aged 16 and over) received a health assessment visit from a registered nurse .A range of anthropometric, physical function, physiological, and biological measures were collected from approximately 20,000 adults, with non-fasting blood samples collected from a subset of 13,130 individuals. Full details of the nurse visits, including eligibility criteria, sampling procedures, and assay methods, have been published elsewhere (University of Essex, 2025). For respondents of the Wave 2 core interview, UK Household Longitudinal Study (UKHLS) participants had nurse visits after their Wave 2 interview; whereas British Household Panel Survey (BHPS) participants had their nurse visits sampled as part of “Wave 3” nurse data. For our anthropometric, cardiovascular, and metabolic outcomes, nurse visits occurred an average of eight months (SD=5) after Wave 2 interviews, with the majority of the nurse visits being categorised as Wave 2 nurse data (75-77%) (Supplement A).

Recognising that relationships between social connections and biological markers can be bidirectional (Gao et al., 2024), but due to data limitations, we focused on cross-sectional analyses with the aim of identifying associations that could be used as the basis for causal directional studies in future research. We took a complete-case approach for our analyses, using social connection exposures and covariate data from Wave 2 and nurse visit data collected across Waves 2/3 (the only timepoint that nurse data has been collected at the time of publication). We excluded participants with missing exposure, covariate, or outcome data, as well as outcomes with undetectable measurements. However, to maximise sample size, we allowed sample sizes to vary across the three types of outcomes assessed, yielding n=16,284 for anthropometric outcomes, n=13,902 for cardiovascular outcomes, and n=10,070 for metabolic blood biomarkers.

For main analyses (see Statistical analyses below), robustness checks adjusting for data collection gaps (in months) between Wave 2 interviews and nurse visits (Waves 2/3) found no tangible coefficient differences across analyses, suggesting that the time gap did not materially alter social connection and health marker associations (Supplement B).

### Outcomes

Eight markers were categorised into three groups: anthropometric markers, including BMI (kg/m^2^) and waist circumference (cm); cardiovascular markers, including pulse rate (bpm), systolic blood pressure (BP) (mmHg), and diastolic BP (mmHg); and metabolic biomarkers, including total cholesterol (mmol/L), triglycerides (mmol/L), and glycated haemoglobin (HbA1c) (mmol/mol). Details of the collection protocols are provided in the Supplement (Table S1). All of the outcome variables were treated as continuous.

As biomarkers can exhibit substantial positive skewness and kurtosis, we examined the distribution of each blood marker for skewness and kurtosis in the final analytical sample (n=10,070). Before log transformation, the skewness and kurtosis values were 2.53 and 14.66 for triglycerides, 4.11 and 31.60 for HbA1c, and 0.41 and 3.30 for total cholesterol. This indicated that triglycerides and HbA1c required a natural log transformation to correct for skewness. After log transformation, the skewness and kurtosis values were 0.30 and 2.99 for triglycerides, and 1.87 and 11.57 for HbA1c. Although the kurtosis for HbA1c remained relatively high, it was retained for analysis; which may be considered a limitation of these analyses.

### Exposures

In the current study, the conceptualisation of our measures was based on a well-established framework for social connections (Holt-Lunstad, 2018; Holt-Lunstad et al., 2017). Due to limitations in the social connections data available within Understanding Society at Wave 2, we were unable to examine all measures of social connections. However, we were able to select one measure for each of the structural, functional, and quality dimensions with the data that was available, acknowledging that these measures are not intended to serve as proxies for each dimension as a whole, but rather represent one example of an indicator within each dimension.

For the structural dimension, we focused on the indicator of living with others. This was measured using household size, which was dichotomised into ‘0’ = living alone and ‘1’ = living together with others (i.e., living with at least one other person).

For the functional dimension, we focused on the indicator of social support. Three questions were asked for each relationship type: family, friends, and partner. The questions were: “How much do they really understand the way you feel about things?”, “How much can you rely on them if you have a serious problem?”, and “How much can you open up to them if you need to talk about your worries?”. Items were rated on a scale from 0 to 3 (0 = not at all, 1 = a little, 2 = somewhat, and 3 = a lot). The scores were summed for each relationship type individually, and the mean score across all three relationship types was then computed to create an average measure of social support (range: 0-9), with higher scores indicating greater social support. Across the three analytical samples, 63.8%-65.4% of participants had all three relationship types, 31.5%-33.2% had two relationship types, and 3.0%-3.2% had one relationship type. Individuals with no reported relationship types were excluded from this measure because they were not asked the social support questions if they had not identified having those relationships. This measure of social support has been used extensively in the literature across different cohort studies (Al-Khouja et al., 2021, p. 1; Iob et al., 2018; Walen & Lachman, 2000, p. 200).

For the quality dimension, we focused on the indicator of relationship strain. This was also assessed across family, friends, and partner relationships. The three questions asked were: “How much do they criticise you?”, “How much do they let you down when you are counting on them?”, and “How much do they get on your nerves?”. These three items were rated on a scale from 0 to 3 (0 = a lot, 1 = somewhat, 2 = a little, and 3 = not at all). The scores were summed for each relationship type individually, and then the mean score across all three relationships was computed to get an average measure of relationship strain (range: 0-9), with higher scores indicating lower relationship strain. Across the three analytical samples, 63.1%-64.8% of participants had all three relationship types, 31.8%-33.5% had two relationships, and 3.4%-3.6% had one relationship. individuals with no reported relationship types were excluded from this measure because they were not asked the relationship strain questions if they had not identified having those relationships. Similar measures of relationship strain have also been previously used in other studies (Chen & Feeley, 2014; Maki, 2020; Stafford et al., 2011).

### Covariates

We identified several potential confounders of the associations between social connections and anthropometric, cardiovascular, and metabolic markers of health. These included sociodemographic factors such as age (in years), sex (female, male), self-reported ethnicity (ethnic minority group, white ethnicity), education level (no qualification, other qualification, GCSE or equivalent, A-level or equivalent, degree or higher), employment status (unemployed, employed), household income (in quintiles), and urbanicity (rural area, urban area).

In analyses examining social connections as the exposure, there is an important balance between under- and over-adjustment. We also considered additional covariates that could either act as confounders or lie on the causal pathway between social connections and health outcomes. Accordingly, these variables were added iteratively in subsequent models as part of additional sensitivity analyses to assess the consistency of findings across different levels of adjustment. These additional covariates included the presence of a disability or long-term health condition (no, yes), smoking status (never, ex-smoker, current smoker), alcohol consumption never or rarely (i.e., never or not at all on the last 12 months), once or twice a month or less (i.e., once or twice a year, once every couple of months, once or twice a month), once or twice a week (i.e., once or twice a week), and more than once or twice a week (e.g., three or four days a week or more), self-reported physical activity (0=no sport at all to 10=very active through sport), and psychological distress (measured using the General Health Questionnaire (GHQ)).

### Statistical analysis

In our main analyses, separate linear regressions were conducted to test the associations between each of the three social connection exposures and each of the cardiovascular, metabolic, and anthropometric outcomes. Model 1 was treated as our main model and adjusted for sociodemographic factors, including age, sex, ethnicity, education level, employment status, household income, and urbanicity, and we also had Model 0 which was unadjusted. Linearity was assessed using scatterplots and assumptions were met. Huber-White robust standard errors were applied to handle homoscedasticity violations. For main analyses (Model 1), to account for multiple testing, p-values were adjusted using the Benjamini–Hochberg procedure, with a false discovery rate of 5%.

In additional analyses, we tested interaction terms to examine whether sex moderated the associations between each social connection measure and each outcome in Model 1. We also ran two sets of supplementary covariate adjustments. Model 2 additionally adjusted for long-term health conditions, smoking status, alcohol consumption, and physical activity. Model 3 further adjusted for psychological distress.

All analyses were conducted using Stata v.18.

## Results

### Sample descriptives

The three analytical samples showed similar descriptives (Table 1). For example, the mean age was approximately 51 years. The participants were relatively equally divided by sex, with 55-56% being female, but were predominantly of white ethnicity (96-97%). The majority of participants lived with other people (84-85%). The mean social support scores were 6.7 (SD=1.6) and the mean relationship strain scores were approximately 7.0 (SD=1.4). The distributions of the outcomes are presented in violin plots (Figure 1). All markers demonstrated relatively normal distributions.

The correlations between all exposures and outcomes are presented in Figure 2. Across the outcomes, the strongest positive correlations were observed between BMI and waist circumference, systolic and diastolic BP, and waist circumference and triglycerides. Across the exposures, living together (our structural indicator of social connections) was weakly correlated with functional and quality aspects (i.e., support and strain). Social support and relationship strain showed a positive correlation, likely due to functional and quality aspects sharing a degree of subjectivity. Overall, the correlations support our approach of treating exposure measures as distinct aspects of social connections.

**Table 1.**
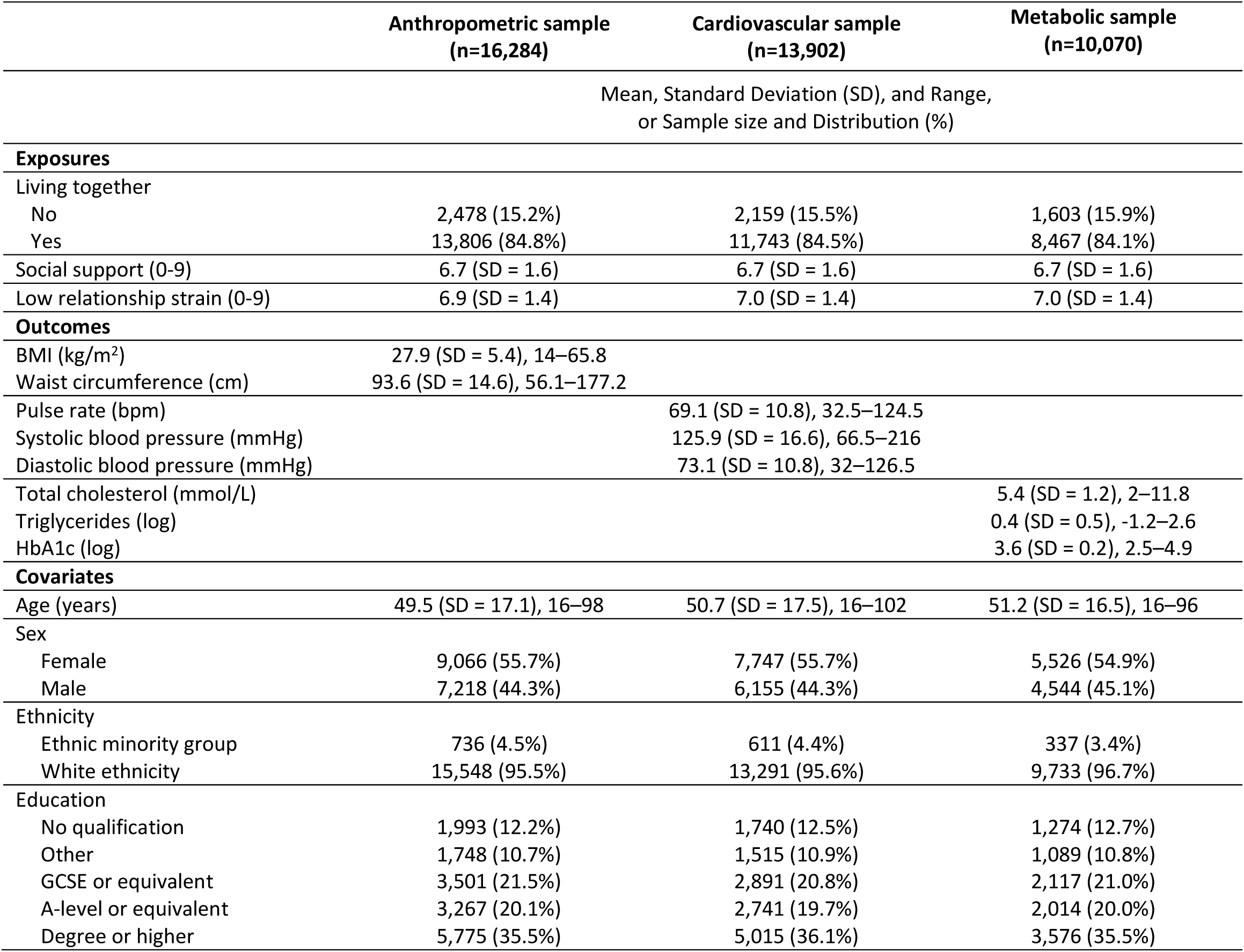
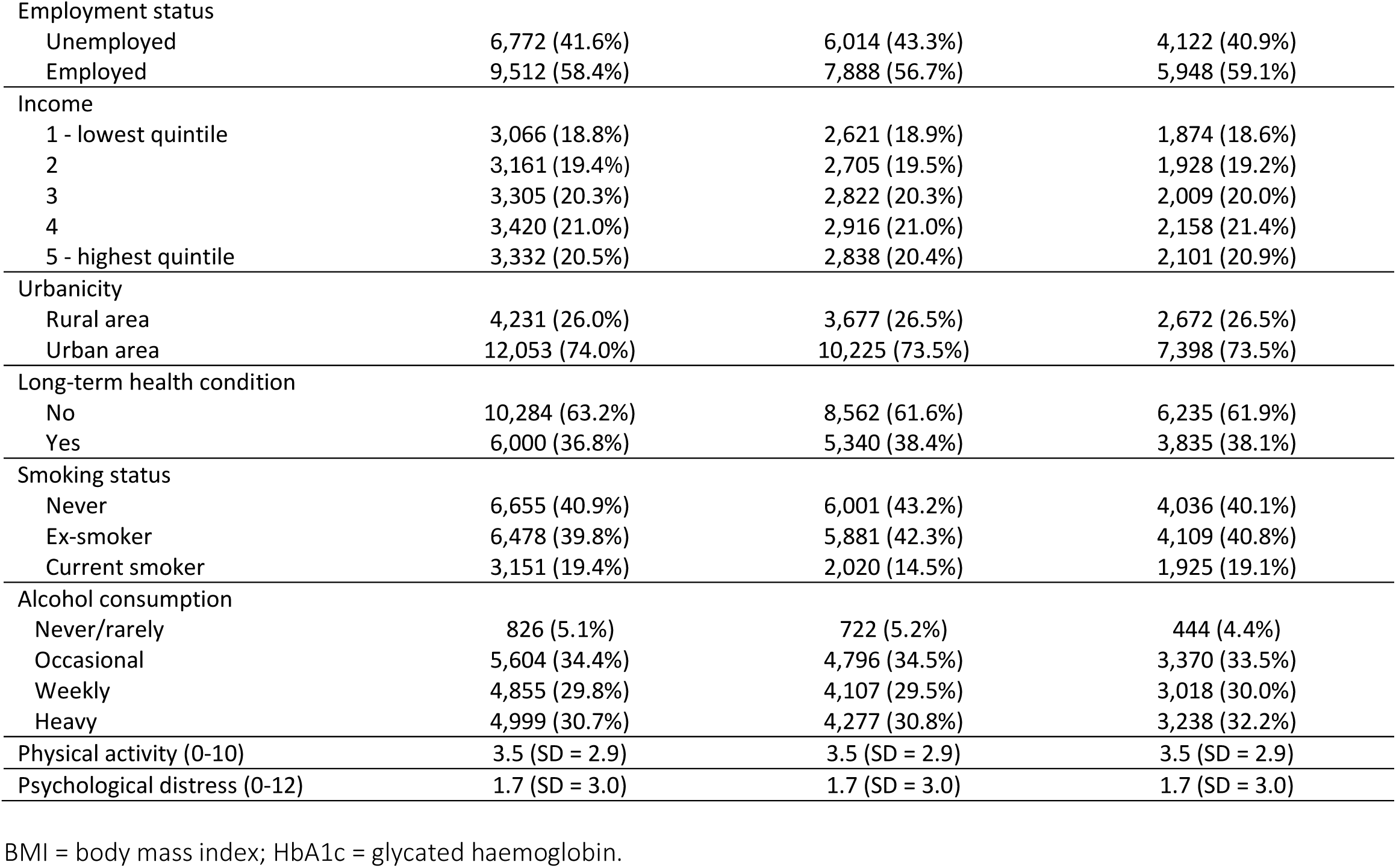
Descriptives of the anthropometric, cardiovascular, and metabolic samples.

**Figure 1.**
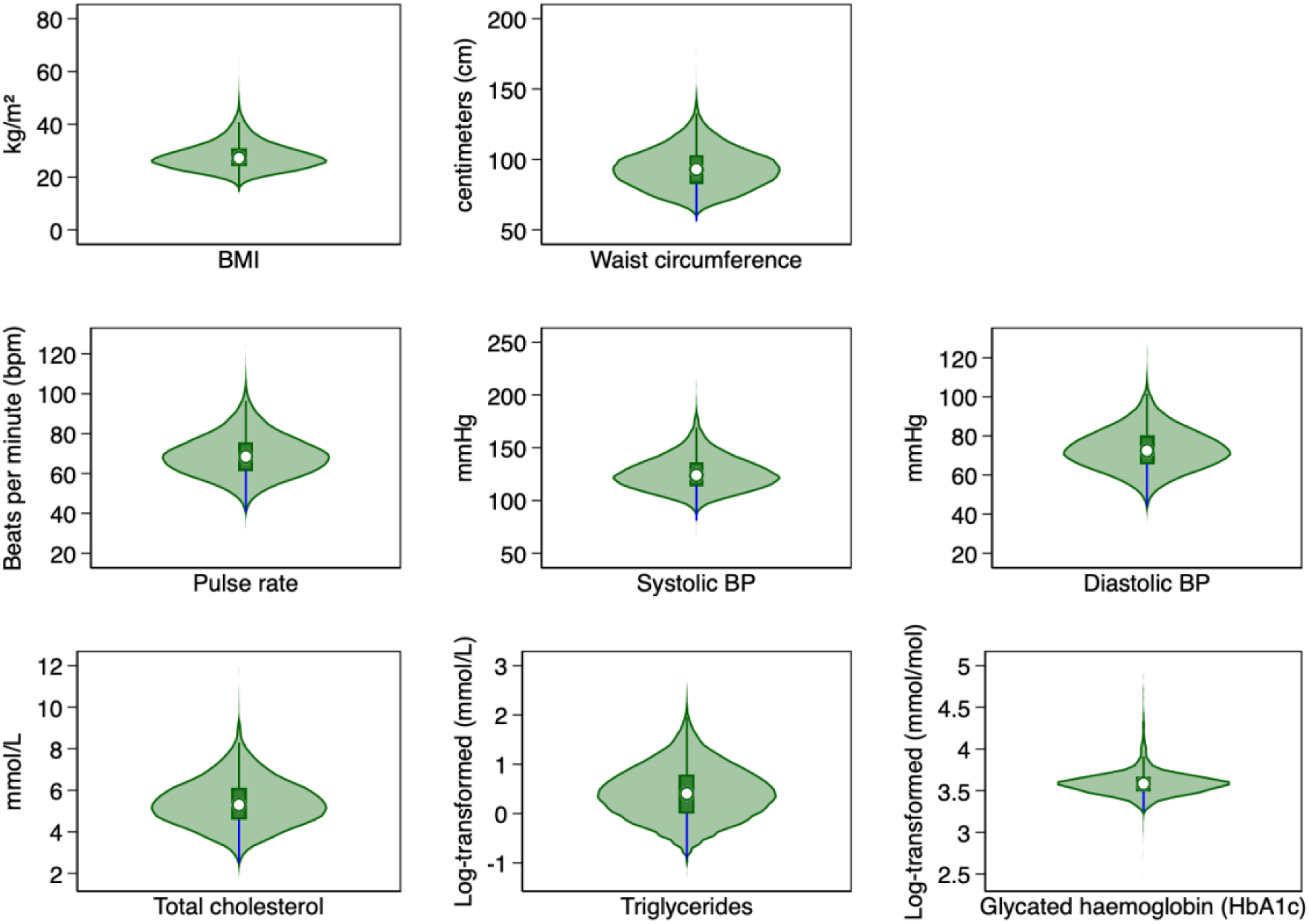
Violin plots of the distribution of outcomes across the anthropometric (n=16,284), cardiovascular (n=13,902), and metabolic (n=10,070) samples. BP = blood pressure.

**Figure 2.**
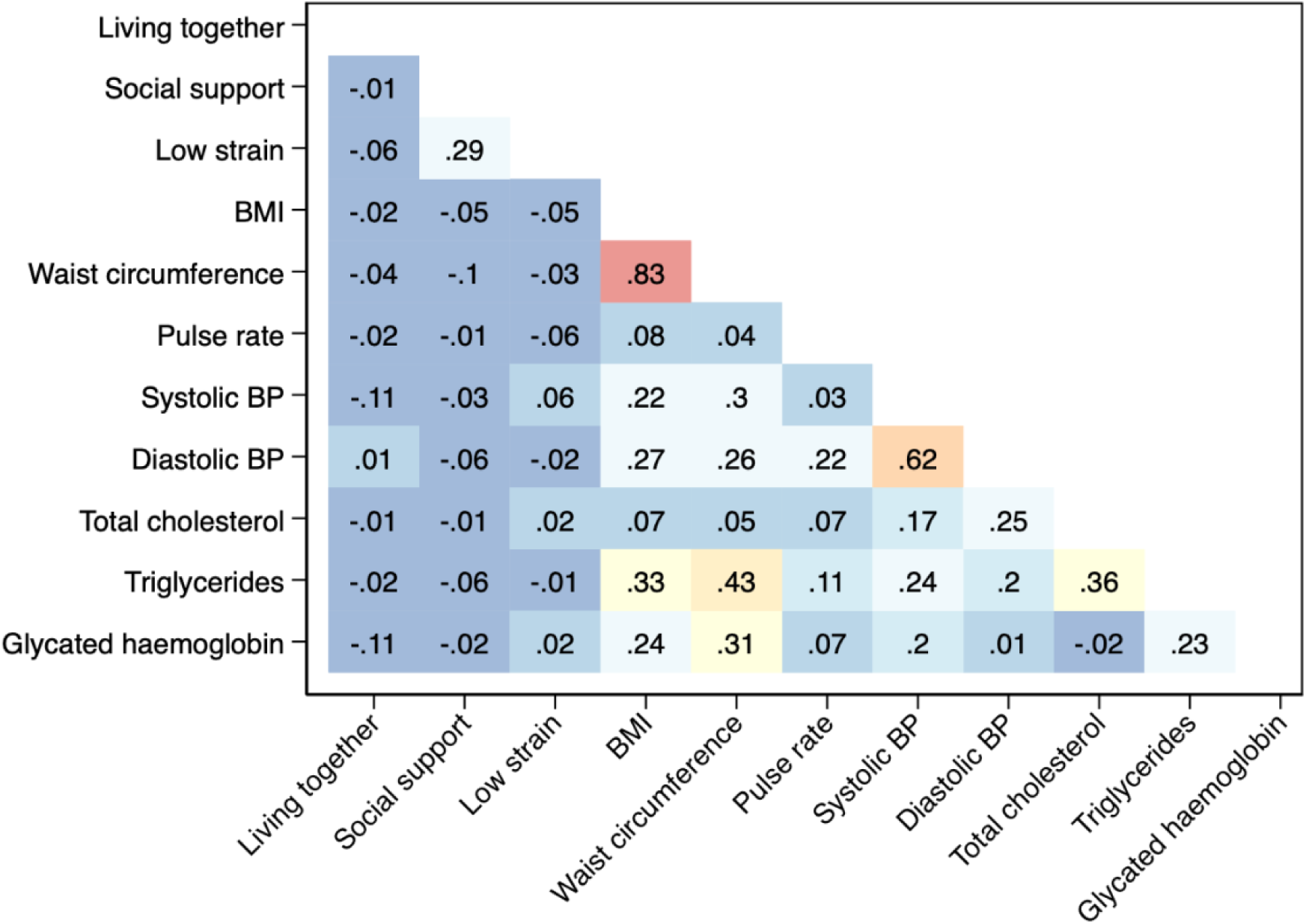
Heat plot of correlations between social connection exposures and anthropometric, cardiovascular, and metabolic markers. The sample size was restricted to complete-cases for all variables in order to test correlations (n=8,084). BMI = Body mass index; BP = blood pressure.

### Main analyses

Findings adjusted for core confounders (Model 1) are presented in Figure 3 and Supplement Tables S2-4.

### Social connections and anthropometric outcomes

Living together with others was associated with higher BMI (B = 0.638, 95%CI = 0.365 to 0.911), whereas greater social support and lower relationship strain were associated with lower BMI (B = -0.165, 95%CI = -0.220 to -0.110; B = -0.249, 95%CI = -0.316 to -0.183).

Similarly, living with others was associated with higher waist circumference (B = 1.597, 95%CI = 0.928 to 2.265), whereas greater social support and lower relationship strain were associated with lower waist circumference (B = -0.459, 95%CI = -0.591 to -0.326; B = -0.624, 95%CI = -0.784 to -0.463).

### Social connections and cardiovascular outcomes

All exposures were related to lower pulse rate: living together with others (B = -0.628, 95%CI = -1.248 to -0.008), greater social support (B = -0.151, 95%CI = -0.265 to -0.036), and lower relationship strain (B = -0.284, 95%CI = -0.419 to -0.148).

Whilst none of the exposures were related to systolic BP, greater social support and lower relationship strain were associated with lower diastolic BP (B = -0.292, 95%CI = -0.407 to -0.178; B = -0.240, 95%CI = -0.378 to -0.102).

### Social connections and metabolic outcomes

Living with others was not associated with total cholesterol. However, greater social support and lower relationship strain were associated with lower levels (B = -0.019, 95%CI = -0.033 to -0.005; B = -0.022, 95%CI = -0.039 to -0.006).

Greater social support and lower relationship strain were associated with lower log-transformed triglyceride levels (B = -0.012, 95%CI = -0.018 to -0.005; B = -0.014, 95% CI = -0.022 to -0.006). This corresponds to approximately 1.2% and 1.4% lower triglyceride levels per unit increase in the exposure, respectively. There was no association for living with others.

Living with others was not associated with HbA1c. However, greater social support and lower relationship strain were associated with lower log-transformed HbA1c levels (B = -0.003, 95%CI = -0.005 to -0.001; B = -0.006, 95% CI = -0.008 to -0.003), equating to approximately 0.3% and 0.6% lower HbA1c levels per unit increase in the exposure, respectively.

After adjustment for multiple testing, all significant results remained except for the association between living together and pulse rate.

**Figure 3.**
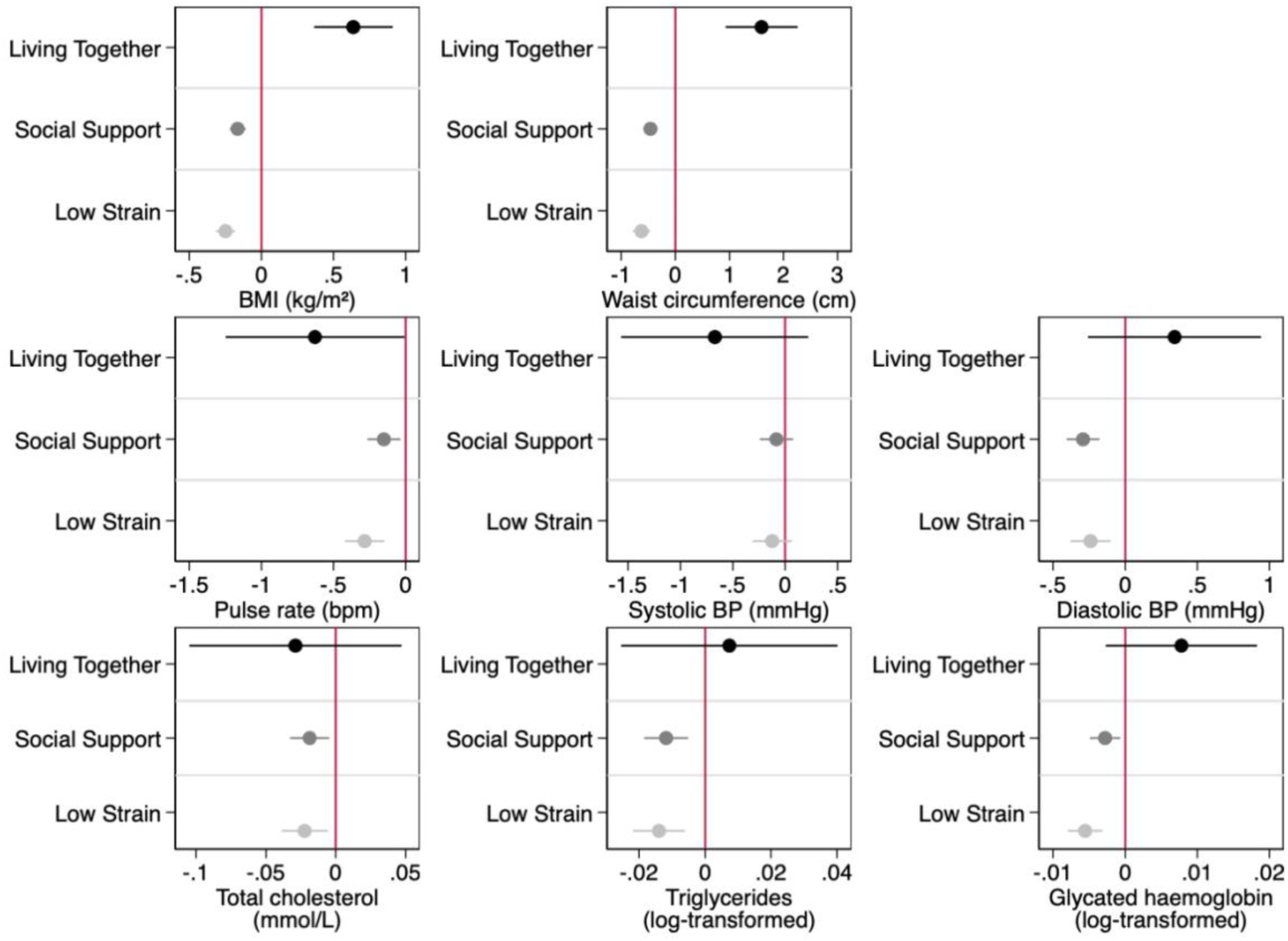
Coefficient plots showing the associations between social connection exposures and anthropometric, cardiovascular, and metabolic outcomes. Model 1 controlled for age, sex, ethnicity, education level, employment status, household income, and urbanicity. The sample sizes were n=16,284 for anthropometric outcomes, n=13,902 for cardiovascular outcomes, and n=10,070 for metabolic outcomes. BP = blood pressure. Graphs present unstandardised beta coefficients and 95% confidence intervals.

### Additional analyses

#### Covariate models

When incorporating additional factors into models that could act as confounders or may lie on the causal pathway, results were maintained across the anthropometric outcomes (BMI and waist circumference), but with some attenuation of coefficient size (Supplement Tables S2-4).

For cardiovascular outcomes, results were maintained for all results, except the association between social support and pulse rate did not hold after additional adjustment for health-related variables.

For metabolic outcomes, results were largely maintained for all results except for attenuation for social support after additional adjustment for health-related variables in relation to hbA1C and for psychological distress in relation to triglycerides.

#### Interactions

When exploring sex interactions (Fig 4, Tables S5-7), there was evidence that the inverse associations between relationship strain and BMI, and between relationship strain and waist circumference, were stronger among females than males.

For cardiovascular outcomes, the inverse association between living together and pulse rate was weaker among females than males. The association between living together and blood pressure differed by sex, with a positive association observed in males and an inverse association in females. The association between relationship strain and systolic blood pressure also differed by sex, with an inverse association observed in males and a positive association observed in females.

For metabolic outcomes, the associations between social support and total cholesterol differed by sex, with an inverse association estimated for males and a positive association for females. The inverse association between relationship strain and total cholesterol was weaker among females than males. Finally, the association between living together and triglycerides differed by sex, with a positive association observed among males and an inverse association estimated among females.

**Figure 4.**
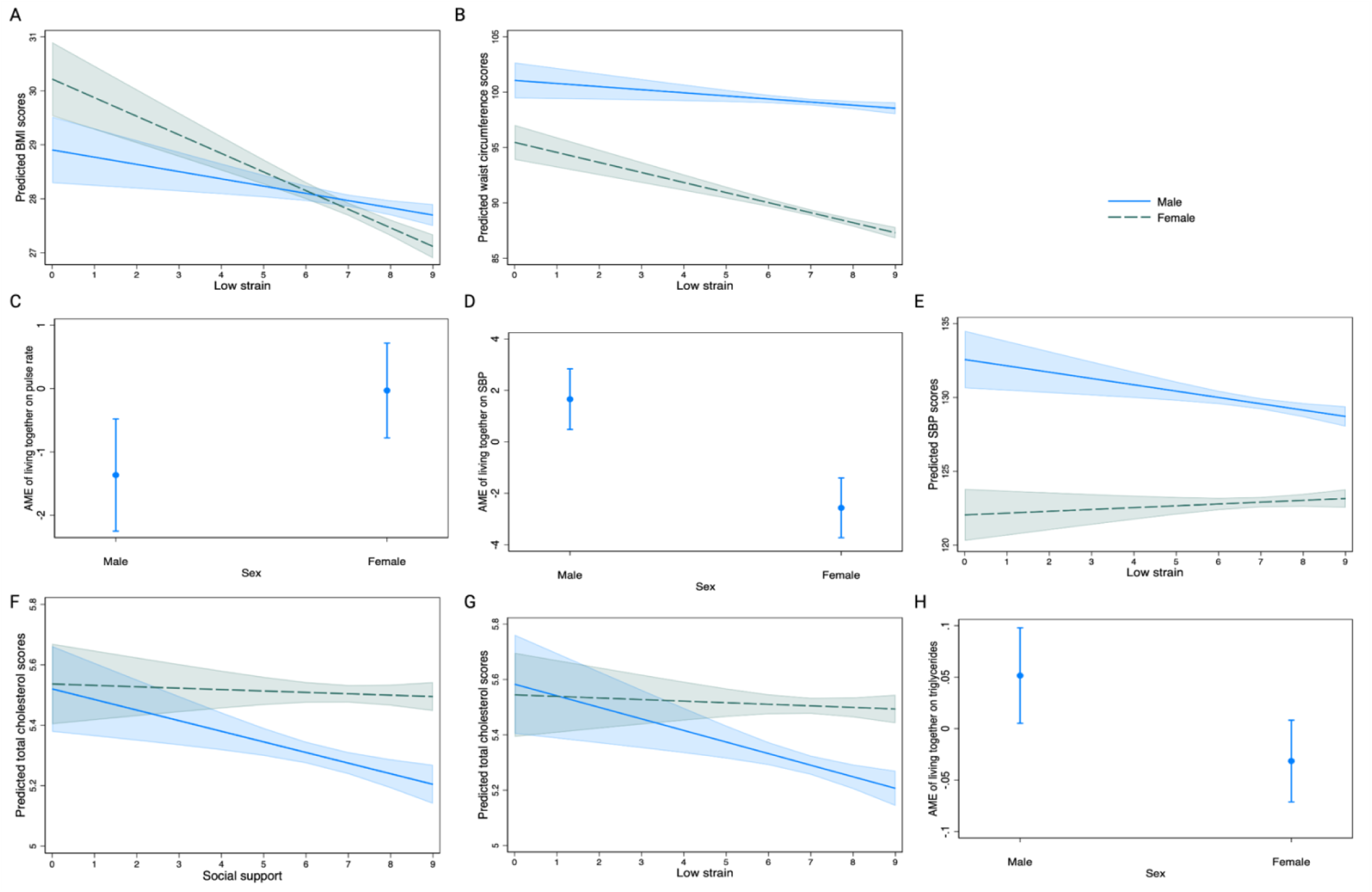
Interaction plots showing the moderating role of sex on the associations between social connections and anthropometric, cardiovascular, and metabolic outcomes. Model 1 controlled for age, sex, ethnicity, education level, employment status, household income, and urbanicity. The sample sizes were n=16,284 for anthropometric outcomes, n=13,902 for cardiovascular outcomes, and n=10,070 for metabolic outcomes. AME = average marginal effect; SBP = systolic blood pressure. Figures C, D, and H present average marginal effects. Figures A, B, E, F, and G present predicted outcomes.

## Discussion

This study investigated how structural, functional, and quality aspects of social connections were associated with anthropometric, cardiovascular, and metabolic markers of health. Living with others was associated with higher BMI and greater waist circumference but a lower pulse rate. Greater social support and lower relationship strain were associated with lower levels across multiple markers, suggesting a protective role of these social connections. All but one finding remained robust after correction for multiple testing, with most associations persisting following further covariate adjustment in additional analyses. There was some evidence of modest moderation by sex. Overall, these findings demonstrate that distinct dimensions of social connections show nuanced associations with anthropometric, cardiovascular, and metabolic markers, contributing to our understanding of the potential biological pathways linking social connections with health.

It is notable that there was a distinction in the associations between living with others and anthropometric outcomes and those between social support and low relationship strain. Previous research has similarly shown that living with others and entering marriage are linked with higher BMI and obesity prevalence (Madani Civi et al., 2024; Yan & Liu, 2023), greater emotional support with flatter BMI and waist -to-hip ratio trajectories (Tymoszuk et al., 2019), and high relationship strain with greater odds of waist circumference increases over time (Kershaw et al., 2014). Shared living arrangements may influence dietary behaviours through greater shared food intake and less individual control over food choices, which may help explain why living with others was associated with higher BMI and waist circumference in our study (Powell et al., 2015). Conversely, social networks can also promote healthier eating behaviours, alongside other health-related behaviours such as increased physical activity and alcohol reduction (Aral & Nicolaides, 2017; Van Den Ende et al., 2024). This may help explain why greater social support and lower relationship strain were associated with more favourable anthropometric outcomes. Our findings also echo previous research demonstrating sex differences in these associations (Hosseini et al., 2021; Hosseini et al., 2020; Tymoszuk et al., 2018). We found that the protective associations between lower relationship strain and lower BMI and waist circumference were stronger among females than males. Having less relationship strain, and therefore potentially less relationship-related stress in one’s relationship, may offer females greater protection against stress-related behaviours such as emotional eating or eating less healthily (Klatzkin et al., 2022; Taylor et al., 2018), thereby contributing to lower BMI and waist circumference. These findings may reflect sex differences in how interpersonal stress and relationship quality are experienced and translated into health-related behaviours, with women potentially being more sensitive to the quality of close relationships, and their influence on adiposity markers. However, evidence for sex differences remains mixed and could also interplay with age. For example, one study only found protective effects of low strain for BMI in older men (Tymoszuk et al., 2019). Together, these findings suggest that different aspects of social connections may have distinct associations with adiposity markers, with structural aspects differing from functional and quality-related dimensions. Importantly, our findings suggest that simply having others around may not confer the same benefits for BMI and waist circumference as having supportive, quality connections.

We also found that greater social support and lower relationship strain were associated with lower diastolic BP. Although some experimental evidence supports these findings for social support (Uchino et al., 2013), recent meta-analyses of experimental studies have reported no consistent associations between social support and BP (Landvatter et al., 2025; Uchino et al., 2022). One possible explanation for the discrepancy between our findings and those of previous experimental studies is the different contexts in which social relationships and BP have been assessed. Previous experimental studies have typically examined ambulatory BP in daily life or BP responses to acute laboratory stressors, often in relatively small samples. In contrast, in the present study, BP was measured during a general nurse visit rather than in direct response to an acute stressor or in relation to in-the-moment reflections on social connections. Instead, social support and relationship strain reflected participants more general perceptions of their relationships, on average 8 months before the BP measurement. However, findings from our analyses need to be replicated and indeed analyses from other cohort studies have generally found inconclusive evidence for associations between social support and BP (Yang et al., 2016). We also found that greater social support and lower relationship strain were associated with lower pulse rate. Across the literature, most studies have assessed heart rate rather than pulse rate, often in relatively small experimental studies examining whether social connections buffer cardiovascular responses to acute stress (O’Riordan & Costello, 2025; Uchino & Garvey, 1997). Therefore, our findings provide additional insight into the potential role of social connections and cardiovascular markers by exploring associations in a large cohort study using measurements obtained during routine health assessments. Nevertheless, given the mixed evidence across the literature, further research is needed to replicate and clarify our findings.

We also observed associations between greater social support and lower relationship strain with lower levels of total cholesterol, triglycerides, and HbA1c. While smaller-scale observational studies have suggested that associations may exist between social connections and some metabolic markers (Edes et al., 2022), epidemiological evidence remains surprisingly sparse. Of the studies that have been conducted, these have generally suggested that social support and relationship strain are not associated with HbA1c (Ford & Robitaille, 2023; Maki, 2020; Yokobayashi et al., 2017). This difference between our HbA1c findings and existing evidence may be because the aforementioned studies used different methods and populations. For example, Ford et al. (2023) found no associations between social support and relationship strain with HbA1c. Although they used identical measures of social support and relationship strain to ours, they focused solely on spouses in older adults and assessed changes over time. Maki (2021) assessed support from friends, family, and spouses in cross-sectional analyses, but only found a direct association between greater social support from friends and lower HbA1c levels in mid-life participants. Yokobayashi et al, (2017) also found that, cross-sectionally, social support (which was measured differently from our study) was not associated with HbA1c in older adults. Therefore, to our knowledge, the associations between social connections and metabolic markers observed in our study present novel findings. Lastly, some of our associations appeared to be moderated by sex. For example, the associations between social support and total cholesterol, and relationship strain and total cholesterol, were stronger among males. One proposed reason could be because the associations between greater social support and its beneficial impacts on health behaviours (e.g., healthier eating, smoking cessation, and more physical activity) and cholesterol screening uptake has been found to be stronger in men than women (Blakoe et al., 2023), with stress potentially having a greater influence on less physical activity among men (Taylor et al., 2018). This may explain why more favourable associations were found in men than women for these metabolic outcomes. However, more research is needed adopting causally informed longitudinal designs, whilst exploring the role of different relationship types, age, and sex.

The current findings should be interpreted in light of several strengths and limitations. The study design was cross-sectional due to data availability. Therefore, there was not the opportunity to explore bidirectional associations by running cross-lagged or other temporal analyses, limiting causal inference. However, analyses were conducted in a large cohort sample, enhancing the generalisability of the findings compared with previous experimental research. We additionally explored associations when considering the time gap between interviews and nurse assessments but found no material changes. However, we do recognise that these different markers vary in their temporal responsiveness to stimuli and variability over time, which affects interpretation. For example, heart rate can change relatively rapidly in response to social exposures, whereas BMI changes more slowly. Consequently, associations between social connections and health markers may reflect processes operating over different timescales. Additionally, these markers are influenced by numerous exposures beyond social connections, and the mechanisms linking social connections to these markers are likely to interplay with a range of other biological processes, as well as psychological, social, and behavioural processes.

We were cautious about over-adjusting our statistical models, as many factors could lie on the causal pathway between social connections and our health markers. Therefore, we focused on a model adjusting for the minimum necessary set of confounders, followed by incremental adjustment for additional covariates to examine whether the observed associations remained consistent. Results remained materially similar even with this further adjustment. Nonetheless, residual confounding remains possible. As only a single measure was used to assess each social connection domain, we acknowledge that this does not fully capture their complexity, so our findings are not intended to be used to extrapolate to other measures within each domain. However, assessing multiple dimensions of social connections, based on an established conceptual framework, is a strength of this study as it allowed comparison of the findings within the same data; something that has been infrequent in past analyses (Hanna et al., 2023; W. Wang et al., 2023). The implications of our findings should also be interpreted in light of their clinical relevance. For instance, lower BMI scores of 0.1-0.2 kg/m^2^, or higher scores of 0.6 kg/m^2^, are not considered clinically significant. Nevertheless, even modest effects may translate into real life changes in health outcomes if occurring as part of broader profiles of biological dysregulation. We therefore encourage future research to assess these relationships longitudinally by investigating how anthropometric and cardiometabolic markers change over time in relation to social connections and examining how trajectories or profiles of social connections influence these markers. Also, given the well-established relationships between social connections and cardiometabolic outcomes, we also encourage studies to test how these markers implicated with glycemic control, lipid metabolism, and adiposity may mediate such relationships, an area of research that is currently sparse.

## Conclusion

Our study has demonstrated a nuanced pattern of association between structural, functional, and quality aspects of social connections and anthropometric, cardiovascular, and metabolic markers of health. These findings have a relevance in light of research linking social connections to cardiometabolic outcomes and highlight promising candidate biological markers for further testing in mediation studies. To further advance this work, we recommend that future studies use a broad panel of social connection measures captured within a single dataset to elucidate specifically which aspects are most relevant to which markers and consider how patterns of social connections across the life-course may differentially relate to biomarkers. We also advocate for the usage of high-throughput molecular biomarker data to enable a transition from a hypothesis-driven approach focused on specific biomarkers to a data-driven approach that could uncover novel biological pathways influenced by social connections. Overall, our findings contribute to the evidence base for the continued consideration of social connections as a public health priority and for social and community-based approaches for population health.

## Data Availability

Data for this study was accessed via the UK Data Service.

https://datacatalogue.ukdataservice.ac.uk/series/series/2000053#access-data

## Acknowledgements

Funding for this research was provided as part of the Understanding Society Fellowship programme, a component of the Study’s Economic and Social Research Council award [ES/S007253/1]. This research was also supported by UK Research and Innovation [MR/Y01068X/1].

## Author contributions

SF, ER, and DF conceptualised and designed the study. ER undertook the preliminary data preparation and data analysis. SF undertook the formal data preparation and data analysis. ER and SF prepared the figures and tables. ER drafted the original manuscript and SF edited and re-drafted the current manuscript. SF, ER, KT, and DF all contributed to writing, and all authors were involved in the editing, reading, and approval of the final manuscript.

## Ethics statement

Ethical approval for the collection of survey data was received from The University of Essex Ethics Committee and biological data collection was granted by the National Research Ethics Service. Consent for the collection of biological data could be withdrawn at any time, and additional written consent was required for invasive procedures (i.e. blood samples). Data for this study was accessed via the UK Data Service.

## Notes

### Competing Interest Statement

The authors have declared no competing interest.

### Author Declarations

Data for this study was accessed via the UK Data Service.

